# ChAdOx1 n-COV-19 Vaccine Side Effects Among Health Care Workers in Trinidad and Tobago

**DOI:** 10.1101/2021.11.09.21264627

**Authors:** Chavin D. Gopaul, Dale Ventour, Davlin Thomas

**Affiliations:** The North Central Regional Health Authority, Trinidad and Tobago, West Indies; The University of the West Indies, St. Augustine, Trinidad and Tobago, West Indies

**Author notes:** Corresponding Author: Dr. Chavin D. Gopaul.

## Abstract

**Background:** The pharmaceutical firms have been lauded for the swift development, trial, approval, and rollout of various Covid-19 vaccines. However, a key issue in the vaccination campaign relates to vaccine hesitancy due to concerns on Covid-19 vaccines safety.

**Method:** A retrospective longitudinal study was carried out via a telephone validated questionnaire. The questionnaire domains included demographic data, medical and COVID-19 related anamneses, and local and systemic side effects 48 hours after receiving the first dose of the vaccine and 48 hours after receiving the second dose of the vaccine.

**Results:** The questionnaire was administered to a sample of 687 healthcare workers (*Male* = 275; *Female* = 412). The results indicated that the incidence of reported fever, body pain, chills, nausea, myalgia, headache, malaise, fatigue and other systemic symptoms declined significantly 48 hours after administration of the second dose compared to the first dose. The Chi-square test and multiple logistics regression results were consistent in demonstrating that younger vaccine recipients were more likely to report fever, body pain, chills, nausea, myalgia, headache, fatigue and other symptoms compared to older vaccine recipients. The multiple logistics regression indicate that female vaccine recipients had greater odds of reporting headache, fatigue, discomfort and less likely to report no symptoms compared to male vaccine recipients, 48 hours after receiving both doses.

**Conclusions:** The findings indicate that on average, vaccine recipients reported fewer number of local and systemic side effects within 48 hours after receiving the second dose compared to 48 hours after receiving the first dose. The findings have implications on public health policy efforts to lower vaccine hesitancy.

## 1.0 Introduction

### 1.1 Background

The swift development, trial, approval, and rollout of Covid-19 vaccines represent a tremendous achievement by pharmaceutical firms and healthcare professionals (1). However, a key concern that explains vaccine hesitancy among the adult population pertains to the fact that there is limited research evidence on the efficacy and safety of the new Covid-19 vaccines, which are currently being administered (2, 3). This apprehension and vaccine hesitancy were worsened by the news that elderly recipients of ChAdOx1 n-COV-19 vaccines in Norway experienced adverse side-effects and, or allergic reactions (3, 4). Initially, reports indicated that some of the recipients of ChAdOx1 n-COV-19 vaccines had experienced vaccine-induced thrombotic thrombocytopenia (VIIT), which forced countries such as the UK to restrict its rollout among the younger population. Therefore, the main issue appears to be the lack of strong research evidence (data) on the safety of the new vaccines, which have been implemented across the globe (1). This paper seeks to present recent evidence on side effects after administrating the ChAdOx1 n-COV-19 vaccines.

The insight based on recent research evidence on the safety of ChAdOx1 n-COV-19 vaccine indicates that the jab is associated with mild local and systemic side-effects (1, 5, 6). The outcome based on a UK study indicates that among the elderly, 80 years and above, a single dose of ChAdOx1 n-COV-19 vaccine was more likely to lower the risk of hospitalization due to the occurrence of local and systemic side-effects within a period of 3-4 weeks after vaccine administration (5). Furthermore, there is also evidence that the recipients of the ChAdOx1 n-COV-19 vaccine experience common systemic and local side effects a few days after administration of the vaccine (1). According to WHO (2021), common side effects associated with the ChAdOx1 n-COV-19 vaccine, include pain and discomfort at the injection site, fatigue, nausea, chills, and muscle pain. However, the likelihood of the stated side effects occurring differ depending on the individual’s age, sex, previous COVID-19 infections, and the type of vaccine administered. Studies in the Caribbean have been limited to discussing patterns of presented symptoms for SARS COV-19 and predictors of ICU admission. Both papers have alluded to the unique characteristics of patients with COVID-19 in Trinidad and Tobago and the greater need for research especially in this region (7, 8).

### 1.2 Aims

The aim of this paper is to examine the safety of the ChAdOx1 n-COV-19 vaccine in terms of its systemic and local side-effects on recipients 48 hours after administration. There are three (3) specific objectives of this research paper. First, the research paper evaluates and compares the safety of the first and second dose ChAdOx1 n-COV-19 vaccines within a period of 48 hours after it has been administered. Secondly, the paper also compares the side effects of the ChAdOx1 n-COV-19 vaccine among persons younger than 40 years and persons older than 40 years within a period of 2 days (48 hours) after administration. The third objective of this paper is to evaluate and compare the safety of the ChAdOx1 n-COV-19 vaccine by age and sex.

## 2.0 Method

The retrospective longitudinal follow up involved collection of data concerning the immediate post-vaccination experience of healthcare workers of a tertiary hospital after receiving their 1^st^ and 2^nd^ dose of the ChadOx1 nCov-19 vaccine between February and May 2021. The eligible participants were the first set of persons to be vaccinated in Trinidad and Tobago. They were classified as the priority group recommended by the Ministry of Health of Trinidad and Tobago. The survey was done via a telephone interview, with a 7-item questionnaire, which inquired about the post-vaccination symptoms experienced within 48 hours after receiving the 1^st^ and 2^nd^ dose of the vaccine and any pre-vaccination COVID-19 infection history. These follow up calls were done by the Liaison Unit attached to the tertiary hospital. The department obtained feedback regarding the side-effects during the post-vaccination period. A member of the unit entered the de-identified data into a study-specific database. This anonymous data was then solicited through the North Central Regional Health Authority via the Ethics Committee.

The telephone-administered questionnaire consisted of three sections. The first included demographic data (age, sex) and COVID-19 infection history, the second asked for oral verification that the first dose of the ChAdOx1 n-COV-19 vaccine was received and the side-effects experienced within the first forty-eight (48) hours after the first dose, while the third asked for oral verification that the second dose of the ChAdOx1 n-COV-19 vaccine was received and the side-effects experienced within the first forty-eight (48) hours after the second dose. The healthcare workers were asked to orally select the side-effects they experienced from a preset list including local (discomfort at injection site) and systemic side-effects (fever, body pains, chills, nausea, myalgia, headaches, malaise, fatigue and other). This list was compiled from the authors’ judgment based on the relevant literature.

Participation in this study was voluntary and the health care workers received no sort of financial renumeration in order to reduce the risk of response bias.

## 3.0 Results

### 3.1 Statistical Data Analysis

To examine and compare the safety of the ChAdOx1 n-COV-19 vaccine 48 hours after the first and the second dose, descriptive statistical data analysis was first conducted. Specifically, the descriptive statistical analysis entailed presenting the frequency of occurrence of the systemic and local side effects 48 hours after the first and second dose.

All analyses were performed using IBM SPSS V.21 software version. Non-parametric inferential statistical analysis using Chi-square test was also conducted in SPSS statistical package to establish the extent to which the variation in systemic and local side-effects by dose (48 hours after first and second dose) as well as by age (≤40 years group and >40 years group) is significant. The significance of the difference in systemic and local side effects by dose and age group was assessed at the 1%, 5% and 10% significance levels. The ANOVA test was conducted to establish whether the difference in the reported number of local and systemic side-effects, 48 hours after the first and 48 hours after the second dose was statistically significant.

A logistics regression analysis was also conducted to establish the effect of age and gender on the participants’ likelihood of reporting local and systemic side-effect. A *p*-value of less than .05 and a confidence interval of 95% was considered statistically significant.

### 3.2 Demographic Characteristics

The study included a total of 779 health care workers although only 687 (88.2%) were able to be contacted, 92 (10.2%) respondents were unreachable. A summary on the demographic characteristics of the population is presented in Table 1 below. The participants’ demographic profile indicates that 275 (40%) of the study population were male respondents while majority of them, 412 (60%) were female respondents. In terms of the age profile, 492 (71.6%) of the study population consisted of the young participants aged less than 40 years and 21-29 years consisting of 227 (33%) of the population. The older participants aged above 40 years consisted of those within 41-49 years, 118 (17.2%), 51-59 years, 59 (8.6%), 61-69 years, 14 (2.0%), and 70 years and above were 4 (0.6%). The summary statistics in terms of previous Covid-19 history indicates that only two (n = 2) participants had a prior positive case of Covid-19 infections while majority of them [685 (99.7%)] did not report any previous Covid-19 history.

**Table 1:** Demographic Characteristics of the Study Population and Prevalence of Systemic and Local Adverse Side-Effects.

| | ChAdOx1 n-COV-19<br>First Dose (After 48 Hours) | ChAdOx1 n-COV-19<br>Second Dose (After 48 Hours) | Significance<br>($p$-value) |
| --- | --- | --- | --- |
| **Sex:** |  |  |  |
| Male | 275 (40.0%) | 275 (40.0%) |  |
| Female | 412 (60.0%) | 412 (60.0%) |  |
| **Age:** |  |  |  |
| 18-20 Years | 2 (0.3%) | 2 (0.3%) |  |
| 21-29 Years | 227 (33.0%) | 227 (33.0%) |  |
| 31-39 Years | 263 (38.3%) | 263 (38.3%) |  |
| 41-49 Years | 118 (17.2%) | 118 (17.2%) |  |
| 51-59 Years | 59 (8.6%) | 59 (8.6%) |  |
| 61-69 Years | 14 (2.0%) | 14 (2.0%) |  |
| $\geq 70$ Years | 4 (0.6%) | 4 (0.6%) | |
| **Previous Covid-19 History: Yes** | 2 (0.3%) | 2 (0.3%) |  |
| **Systemic Side-Effects:** |  |  |  |
| Fever*** | 386 (56.3%) | 103 (15.0%) | $< .0001$ |
| Body Pain*** | 314 (45.8%) | 99 (14.4%) | $< .0001$ |
| Chills*** | 239 (34.8%) | 33 (4.8%) | $< .0001$ |
| Nausea*** | 70 (10.2%) | 33 (4.8%) | $< .0001$ |
| Myalgia*** | 100 (14.6%) | 23 (3.4%) | $< .0001$ |
| Headache*** | 234 (34.1%) | 110 (16.0%) | < .0001 |
| Malaise*** | 105 (15.3%) | 27 (3.9%) | < .0001 |
| Fatigue*** | 207 (30.2%) | 143 (20.9%) | < .0001 |
| Others*** | 145 (21.1%) | 86 (12.5%) | < .0001 |
| **Local Side-Effects:** |  |  |  |
| Discomfort at Site*** | 176 (25.7%) | 199 (29.0%) | < .0001 |
| **No Symptoms\*\*\*** | 100 (14.6%) | 276 (40.2%) | < .0001 |
| **Number of Side-effects** | **3.03 ± 1.91** | **1.65 ± 1.11** | **< .0001** |

p\*\*\* < 0.0001(\*\*\*Significant at 0.01% threshold level). Chi-square test was used to compare the difference in the prevalence of systemic and local side effects 48 hours after the first and the second dose with a significance level of <.0001 (Exhibit 4, Appendix). ANOVA test was used to compare the difference between the number of side-effects 48 hours after the first dose and the second dose with a significance level of <.0001 (Exhibit 5, Appendix).

### 3.3 Incidences of Vaccine Side-Effect by Dose, Age, Sex, and Previous Covid-19 History

Figure 1 shows the reported adverse-side effects within 48 hours after the first and second dose of ChAdOx1 n-COV-19 vaccine. The insight based on the analysis of Figure 1 (Panel A) indicates that there were significantly lower reported cases of systemic symptoms 48 hours after the second dose compared to the systemic side-effects 48 hours after the first dose. Specifically, as compared to the second dose of ChAdOx1 n-COV-19 vaccine, the highest side-effect in the first dose was fever [386 (56.3%)], followed by body pain [314(45.8%)]. The reported incidences of fever declined from 386 (56.3%) (48 hours after first dose) to 103 (15.0%) (48 hours after the second dose). Similarly, the reported prevalence of body pain decreased from 314 (45.8%) (48 hours after the first dose) to 99 (14.4%) (48 hours after the second dose). However, the lowest variation in side-effects by the ChAdOx1 n-COV-19 vaccine dose was noted in nausea where the reported incidences declined from 70 (10.2%) (48 hours after the first dose) to 33 (4.8%) (48 hours after the second dose).

**Figure 1:**
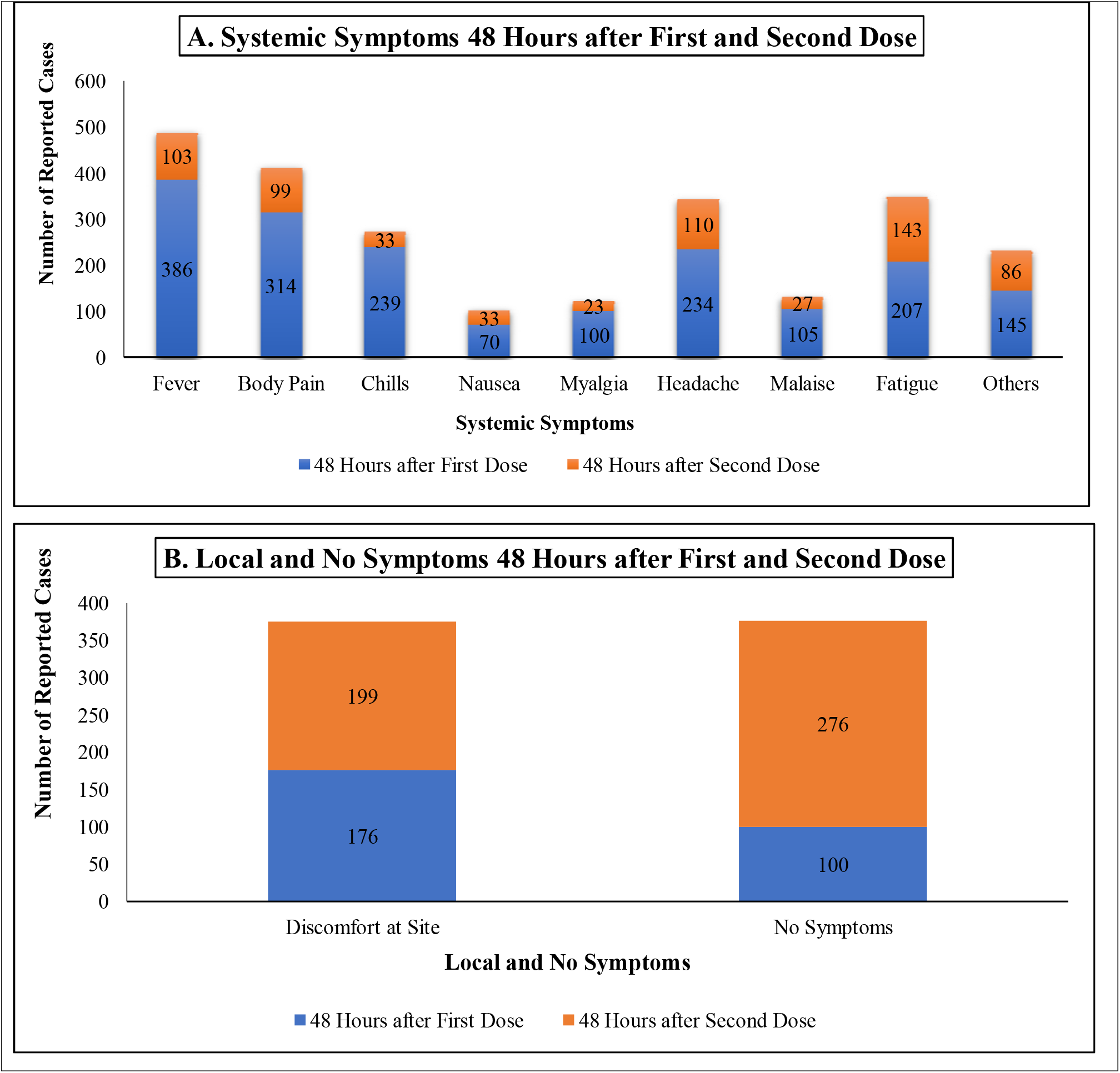
Reported Systemic and Local Side-Effects 48 Hours after First and Second Dose

Figure 1 (Panel B) shows that the number of local symptoms (discomfort at site) were higher 48 hours after the second dose [199 (29.0%)] compared to the reported cases 48 hours after the first dose [176 (25.7%)]. Furthermore, the reported cases of ‘no symptoms’ increased significantly from 100 (14.6%) (48 hours after the first dose) to 276 (40.2%) (48 hours after the second dose).

#### 3.3.1 Analysis of the ChAdOx1 n-COV-19 Vaccine Side-Effects in First and Second Doses Systemic Side-Effects

The results of the descriptive and non-parametric Chi-square test, which were presented for all age groups and both sexes depicted in Table 1 show that a total of 386 (56.3%) reported experiencing fever 48 hours after the first dose of ChAdOx1 n-COV-19 vaccine while only 103 (15.0%) reported feeling fever 48 hours after the second dose. The decrease in the prevalence of fever was statistically significant across the first and the second dose of ChAdOx1 n-COV-19 vaccine (ρ <.0001). Similarly, the results also show that a total of 314 (45.8%) and 99 (14.4%) of the respondents reported experiencing body pain 48 hours after the first and second dose of ChAdOx1 n-COV-19 vaccine respectively. In this case, the decrease in body pain (31.4%; ρ <.0001) after the second dose was statistically significant. A summary of the findings also shows that 239 (34.8%) of the participants reported feeling chills 48 hours after receiving the first dose while 33(4.8%) of the subjects reported experiencing chills, 48 hours after administration of the ChAdOx1 n-COV-19 second dose. The decrease in the frequency of chills symptoms (30%; ρ <.0001) between the first and the second dose of the ChAdOx1 n-COV-19 vaccine is statistically significant. In terms of nausea, 70 (10.2%) of the participants experienced the symptoms 48 hours after the first dose while 33 (4.8%) of the participants reported experiencing nausea side-effects, 2 days after the second dose. Similarly, in this case, the decrease in nausea symptoms (5.4%; ρ <.0001) was statistically significant between the first and the second dose.

The descriptive results also indicate that 100 (14.6%) and 23 (3.4%) of the respondents reported experiencing myalgia symptoms, 48 hours after administration of the first dose and second dose respectively. The decreased incidence of the myalgia symptoms (11.2%; ρ <.0001) was statistically significant. Furthermore, the number of participants who reported feeling headache after the first dose [234 (34.1%)] decreased significantly to 110 (16.0%) 48 hours after administration of the second dose of ChAdOx1 n-COV-19 vaccine. The implication is that the decrease in the reported symptoms of headache was statistically significant (18.1%; ρ <.0001). Similarly, the number of participants who reported feeling malaise [27 (3.9%)] 48 hours after receiving the second dose was significantly lower compared to the number of respondents who reported feeling malaise [105 (15.3%)] 48 hours after administration of the first dose. The decline in the reported symptoms of malaise (11.4%; ρ <.0001) between 48 hours after the first dose and the second dose was statistically significant. In terms of fatigue, the number of participants who reported feeling tired 48 hours after the first dose [207 (30.2%)] was also significantly higher compared to the number of participants [143 (20.9%)] who experienced fatigue within 48 hours after receiving the second dose (ρ <.0001). The occurrence of the other reported symptoms was also significantly lower (8.6% decrease) 48 hours after receiving the second dose [86 (12.5%)] compared to the first dose [145 (21.1%)] (ρ <.0001).

##### Local Side-Effects

The outcome of the descriptive and non-parametric chi-square test presented in Table 1 shows that there was a statistically significant difference in the prevalence of the local side-effects 48 hours after administration of the first and second dose of the ChAdOx1 n-COV-19 vaccine (ρ <.0001). Specifically, the chi-square test results indicate that while 176 (25.7%) of the participants reported feeling discomfort at the site of injection 48 hours after the first dose, the increase in reported symptoms after the second dose [199 (29.0%)] was statistically significant (3.3%; ρ <.0001). This means that the number of participants who reported local side-effects (i.e., discomfort at site) was also significantly different 48 hours after administration of the first and second dose of the ChAdOx1 n-COV-19 vaccine. Lastly, ‘no reported symptoms’ also increased significantly 48 hours after the second dose [276 (40.2%)] compared to the reported no symptoms 48 hours after the first dose [100 (14.6%)] (ρ <.0001). The implication is that ‘no symptoms’ increased significantly 48 hours after administration of the second dose compared to 48 hours after administration of the first dose of the ChAdOx1 n-COV-19 vaccine.

The ANOVA test indicated that on average, the number of reported side-effects declined from 3.03 (*SD* = 1.91) 48 hours after the first dose to 1.65 (*SD* = 1.11) 48 hours after administration of the second dose (ρ <.0001). This means that on average, ChAdOx1 n-COV-19 vaccine recipients had less than two (1.65) reported local and systemic side-effect 48 hours after the second dose compared to the three (3.03) reported after being given the first dose.

#### 3.3.2 Analysis of ChAdOx1 n-COV-19 Side-Effects among Younger and Older Recipients

The results in Table 2 shows that the two participants who had prior Covid-19 infections were aged below 40 years. The implication is that none of the older respondents (> 40 years) reported a positive Covid-19 infection. However, due to the small number of participants who reported prior Covid-19 infections, the Chi-square test results show that there is no statistically significant difference in the Covid-19 anamneses between the younger participants (≤40 years) and the older participants (>40 years).

**Table 2:** Prevalence of Systemic and Local Side-Effects for ChAdOx1 n-COV-19 Vaccine Among Older and Younger Recipients.

| | $\leq 40$ Years | $> 40$ Years | Total | Sig ($p$-value) |
| --- | --- | --- | --- | --- |
| Number of Doses: |  |  |  |  |
| One Dose | 491 (71.6%) | 195 (28.4%) | 686 (100.0%) | $<.0001$ |
| Two Doses | 491 (71.6%) | 195 (28.4%) | 686 (100.0%) |  |
| Previous Covid-19 History: Yes | 2 (0.3%) | 0 (0.0%) | 2 (0.3%) | 0.373 |
| Systemic Side-Effects: |  |  |  |  |
| Fever | 80 (11.7%) | 23 (3.4%) | 103 (15.0%) | 0.137 |
| Body Pain | 75 (10.4%) | 24 (3.5%) | 99 (14.4%) | 0.318 |
| Chills | 24 (3.5%) | 9 (1.3%) | 33 (4.8%) | 0.880 |
| Nausea | 22 (3.2%) | 11 (1.6%) | 33 (4.8%) | 0.522 |
| Myalgia | 18 (2.6%) | 5 (0.7%) | 23 (3.4%) | 0.470 |
| Headache | 81 (11.8%) | 29 (4.2%) | 110 (16.0%) | 0.601 |
| Malaise | 20 (2.9%) | 7 (1.0%) | 27 (3.9%) | 0.769 |
| Fatigue*** | 117 (17.1%) | 26 (3.8%) | 143 (20.9%) | 0.002 |
| Others | 57 (8.3%) | 29 (4.2%) | 86 (12.5%) | 0.244 |
| Local Side-Effects: |  |  |  |  |
| Discomfort at Site* | 152 (22.2%) | 47 (6.9%) | 199 (29.0%) | 0.074 |
| No Symptoms*** | 174 (25.4%) | 102 (14.9%) | 276 (40.2%) | $<.0001$ |

$p^{***} < 0.0001$ (\*\*\*Significant at 0.01% threshold level); $p^* < 0.1$ (\*Significant at 10% threshold level)

The results of the descriptive and Chi-square test presented in Table 2 indicate that there was no statistically significant difference in the reported systemic and local side-effects in younger (≤ 40-year-old group) and older (> 40-year-old group) ChAdOx1 n-COV-19 vaccine recipients based on the previous Covid-19 history (ρ = 0.373). However, based on the Chi-square test findings, the difference in the reported fatigue symptoms between the ≤ 40-year-old group [117 (17.1%)] and the > 40-year-old group [26 (3.8%)] was statistically significant (ρ = 0.002). The non-parametric inferential results based on the Chi-square test also indicate that the ≤ 40-year-old group [152 (22.2%)] reported significantly higher frequency of occurrence of discomfort at the site of injection compared to the > 40-year-old group [47 (6.9%)] (ρ = 0.074) when assessed at the 10% significance level. However, more younger participants, ≤ 40 years [174 (25.4%)] reported no symptoms compared to the older participants, > 40 years [102 (14.9%)] (ρ<.0001) Therefore, the conclusion based on the inferential Chi-square test findings is that more younger individuals experienced systemic side-effects (fatigue) and local side-effects (discomfort) compared to the older individuals.

#### 3.3.3 Effect of Age and Gender on ChAdOx1 n-COV-19 Systemic and Local Side-Effects

This subsection of the results presents the outcome of the multiple logistic regression analysis to estimate the effect of age and gender on vaccine recipients’ likelihood of experiencing systemic and local side-effects. Table 3-5 (First dose panel) show the multiple logistic regression results to ascertain the effect of age and gender on local and systemic side-effects 48 hours after the first dose. The results indicate that younger age (≤40 years) has a significant higher odd on the likelihood of vaccine recipients reporting systemic and local side-effects 48 hours after administration of the first dose. Specifically, younger individuals (≤ 40 years) were 2.60 times (CI 95%: 1.85 - 3.66) likely to report fever and 2.50 times (CI 95%: 1.76 - 3.57) likely to complain of body pain 48 hours after administration of the first dose compared to older vaccine recipients (> 40 years). Similarly, younger vaccine recipients (≤ 40 years) were 2.03 times (CI 95%: 1.39 - 2.96) likely to report chills and 2.05 times (CI 95%: 1.07 - 3.92) likely to experience nausea symptoms 48 hours after the first dose when assessed against the older vaccine recipients (> 40 years). The vaccine recipients who are younger than 40 years were 2.51 times (CI 95%: 1.41 - 4.47) likely to report myalgia and 1.63 times (CI 95%: 1.12 - 2.35) likely to complain of headache 48 hours after receiving the first dose compared to older recipients who are aged above 40 years. The multiple logistic regression results also indicate that younger vaccine recipients (≤ 40 years) were 1.85 times (CI 95%: 1.25 - 2.73) likely to report fatigue and 1.45 times (CI 95%: 0.94 - 2.23) likely to experience other symptoms 48 hours after administration of the first dose compared to older vaccine recipients (> 40 years). In addition, older vaccine recipients aged above 40 years had a higher odds ratio of 2.27 times (CI 95%: 1.47 - 3.52) for reporting no symptoms 48 hours after administration of the first dose compared to the younger vaccine recipients (≤ 40 years).

**Table 3:** Multiple Logistic Regression: Effect of Age and Gender on Fever, Body Pain, Chills and Nausea Side-Effects.

| | | | Fever<br>OR [95% CI] | Sig.<br>ρ-value | Body Pain<br>OR [95% CI] | Sig.<br>ρ-value | Chills<br>OR [95% CI] | Sig.<br>ρ-value | Nausea<br>OR [95% CI] | Sig.<br>ρ-val |
| --- | --- | --- | --- | --- | --- | --- | --- | --- | --- | --- |
| First Dose | Age: | $\leq 40$ Yrs<br>(vs. $> 40$ Yrs) | 2.60 [1.85 - 3.66] | $< .0001$ | 2.50 [1.76 - 3.57] | $< .0001$ | 2.03 [1.39 - 2.96] | $< .0001$ | 2.05 [1.07 - 3.92] | 0.03 |
|  | Gender: | Female<br>(vs. Male) | 1.04 [0.76 - 1.43] | 0.804 | 1.46 [1.06 - 2.00] | 0.019 | 1.59 [1.14 - 2.22] | 0.006 | 2.46 [1.37 - 4.39] | 0.00 |
| Second Dose | Age: | $\leq 40$ Yrs<br>(vs. $> 40$ Yrs) | 1.45 [0.89 - 2.39] | 0.140 | 1.28 [0.78 - 2.10] | 0.321 | 1.06 [0.48 - 2.33] | 0.881 | 0.78 [0.37 - 1.65] | 0.51 |
|  | Gender: | Female<br>(vs. Male) | 1.36 [0.87 - 2.11] | 0.173 | 1.20 [0.77 - 1.87] | 0.417 | 1.03 [0.50 - 2.11] | 0.934 | 1.84 [0.84 - 4.01] | 0.12 |

**Table 4:** Multiple Logistic Regression: Effect of Age and Gender on Myalgia, Headache, Malaise and Fatigue Side-Effects.

| | | | Myalgia<br>OR [95% CI] | Sig.<br>ρ-value | Headache<br>OR [95% CI] | Sig.<br>ρ-value | Malaise<br>OR [95% CI] | Sig.<br>ρ-value | Fatigue<br>OR [95% CI] | Sig.<br>ρ-val |
| --- | --- | --- | --- | --- | --- | --- | --- | --- | --- | --- |
| First Dose | Age: | $\leq 40$ Yrs<br>(vs. $> 40$ Yrs) | 2.51 [1.41 - 4.47] | 0.002 | 1.63 [1.12 - 2.35] | 0.010 | 1.11 [0.70 - 1.77] | 0.663 | 1.85 [1.25 - 2.73] | 0.00 |
|  | **Gender:** | Female<br>(vs. Male) | 1.22 [0.79 - 1.91] | 0.373 | **1.74 [1.24 - 2.42]** | **0.001** | 0.96 [0.63 - 1.46] | 0.842 | **1.51 [1.07 - 2.13]** | **0.01** |
| **Second Dose** | **Age:** | ≤ 40 Yrs<br>(vs. > 40 Yrs) | 1.45 [0.53 - 3.95] | 0.472 | 1.13 [0.71 - 1.79] | 0.608 | 1.14 [0.47 - 2.74] | 0.772 | **2.06 [1.29 - 3.28]** | **0.00** |
|  | **Gender:** | Female<br>(vs. Male) | 1.04 [0.44 - 2.44] | 0.927 | **1.69 [1.09 - 2.62]** | **0.019** | 1.35 [0.60 - 3.06] | 0.467 | **2.16 [1.43 - 3.26]** | **< .000** |

**Table 5:** Multiple Logistic Regression: Effect of Age and Gender on Discomfort at Site, Other Symptoms and No Symptoms Side-Effects.

|  |  |  | Discomfort<br>OR [95% CI] | Sig.<br>p-value | Other Symptoms<br>OR [95% CI] | Sig.<br>p-value | No Symptoms<br>OR [95% CI] | Sig.<br>p-value |
| --- | --- | --- | --- | --- | --- | --- | --- | --- |
| **First Dose** | **Age:** | ≤ 40 Yrs<br>(vs. > 40 Yrs) | 1.04 [0.71 - 1.52] | 0.850 | **1.45 [0.94 - 2.23]** | **0.090** | **2.27 [1.47 - 3.52]** | **< .0001** |
|  | **Gender:** | Female<br>(vs. Male) | **1.41 [0.99 - 2.01]** | **0.060** | 1.16 [0.80 - 1.70] | 0.436 | **1.61 [1.05 - 2.48]** | **0.029** |
| **Second Dose** | **Age:** | ≤ 40 Yrs<br>(vs. > 40 Yrs) | **1.41 [0.97 - 2.07]** | **0.076** | 0.75 [0.47 - 1.22] | 0.246 | **2.01 [1.44 - 2.83]** | **< .0001** |
|  | **Gender:** | Female<br>(vs. Male) | **1.52 [1.07 - 2.14]** | **0.019** | 0.92 [0.58 - 1.46] | 0.724 | **1.68 [1.23 - 2.31]** | **0.001** |

The outcome of the multiple logistics regression analysis presented in Table 3-5 (First dose panel) indicate that gender has a significant positive effect on the likelihood of ChAdOx1 n-COV-19 vaccine recipients reporting systemic and local side-effects 48 hours after the first dose. Specifically, the multiple logistic regression results show that female vaccine recipients were 1.46 times (CI 95%: 1.06 - 2.00) more likely to complain of body pain and 1.59 times (CI 95%: 1.14 - 2.22) more likely to experience chills compared to male vaccine recipients, 48 hours after administration of the first dose. In addition, female vaccine recipients had odds ratio (OR) of 2.46 times (CI 95%: 1.37 - 4.39) for experiencing nausea and were 1.74 times (CI 95%: 1.24 - 2.42) more likely to complain of headache 48 hours after administration of the first dose compared to the male vaccine recipients. The female vaccine recipients were also 1.51 times (CI 95%: 1.07 - 2.13) more likely to report fatigue symptoms and had a higher odds ratio (OR) of 1.41 times (CI 95%: 0.99 - 2.01) for experiencing discomfort at site 48 hours after administration of the first dose compared to male vaccine recipients. In terms of gender, male vaccine recipients were 1.61 times (CI 95%: 1.05 - 2.48) likely to report no symptoms compared to female vaccine recipients 48 hours after administration of the first dose.

Tables 3-5 (Second dose panel) show the multiple logistic regression results to estimate the effect of age and gender on local and systemic side-effects 48 hours after administration of the second dose of ChAdOx1 n-COV-19 vaccine. 48 hours after receiving the second dose, the multiple logistic regression results show that age has a significant effect on the odds of reporting local and systemic side-effects. The specific findings indicate that younger vaccine recipients (≤ 40 years) were 2.06 times (CI 95%: 1.29 - 3.28) likely to report fatigue and had a higher odds ratio of 1.41 times (CI 95%: 0.97 - 2.07) for experiencing discomfort at site 48 hours after receiving the second dose of the ChAdOx1 n-COV-19 vaccine compared to older vaccine recipients (> 40 years). In addition, the multiple logistic regression results indicate that older vaccine recipients aged 40 years and above had a higher odd of 2.01 times (CI 95%: 1.44 - 2.83) for not reporting any symptoms compared to younger vaccine recipients (≤ 40 years) 48 hours after receiving the second dose.

The results of the multiple logistic regression analysis in Table 3-5 (Second dose panel) also show that gender has a significant positive effect on the likelihood of vaccine recipients reporting headache, fatigue, discomfort at site and no symptoms 48 hours after receiving the second dose of ChAdOx1 n-COV-19 vaccine. Specifically, while female vaccine recipients were 1.69 times (CI 95%: 1.09 - 2.62) more likely to complain of headache, they also had a greater odds ratio (OR) of 2.16 times (CI 95%: 1.43 - 3.26) for experiencing fatigue symptoms 48 hours after receiving the second dose compared to male vaccine recipients. The female vaccine recipients were also 1.52 times (CI 95%: 1.07 - 2.14) more likely to report discomfort at site compared to male vaccine recipients, 48 hours after administration of the second dose of ChAdOx1 n-COV-19 vaccine. Finally, male vaccine recipients were 1.68 times (CI 95%: 1.23 - 2.31) likely to report no symptoms compared to female vaccine recipients 48 hours after administration of the second dose.

## 4.0 Discussion

### 4.1 Discussion of Findings

The study sought to examine the side-effects of the ChAdOx1 n-COV-19 vaccine 48 hours after administration of the first and the second dose. The inferential non-parametric test results based on the Chi-square test analysis reveal that there is a significant difference in the reported systemic symptoms (i.e., fever, body pain, chills, nausea, myalgia, headache, malaise, fatigue, and other systemic symptoms) as well as local symptoms (discomfort at site) 48 hours after administration of the first dose and the second dose. The incidence of reported fever, body pain, chills, nausea, myalgia, headache, malaise, fatigue and other systemic symptoms declined significantly 48 hours after administration of the second dose compared to the reported corresponding systemic symptoms, 48 hours after administration of the first dose. Similarly, using ANOVA test, this study also found that the reported number of side-effects declined significantly 48 hours after the second dose compared to the first dose. Specifically, vaccine recipients reported on average less than 2 side-effects 48 hours after the second dose compared to the three first the first dose. These results are consistent with previous findings (9) based on a similar study that sought to evaluate the safety of the ChAdOx1 n-COV-19 vaccine. According to the study by Menni et al. (9), the systemic symptoms, including headache, fatigue, chills, diarrhoea, myalgia, nausea and arthralgia were 1.6 times likely to occur in the first dose compared to the second dose, especially among vaccine recipients with prior Covid-19 infections. The study by Bernal et al. (10) also shows that recipients of the first dose of the ChAdOx1 n-COV-19 vaccine had 37% reduced risk of hospital admissions due to the occurrence of local and systemic side-effects. Similarly, in a study to examine the side-effects of the ChAdOx1 n-COV-19 vaccine in Jordan, (11) finds that only 32.2% of the vaccine recipients reported mild systemic side-effects such as headache, fever, chills, and myalgia. On the other hand, 33.5% of the ChAdOx1 n-COV-19 vaccine recipients experienced moderate local side-effects such as pain at the site of injection after administration of the ChAdOx1 n-COV-19 vaccine However, these findings contrast the outcome based on a systematic review study by (12). Based on the findings from the study, which sought to evaluate the safety of the two-dose regimen of ChAdOx1 n-COV-19, there was no significant variation in the reported systemic and local side-effects after the first and the second dose of the ChAdOx1 n-COV-19 vaccine (12). In other studies, (13, 14), it was also found that there is no significant variation in the reported side-effects (headache, fever, myalgia, chills, pain at site, shiver, and nausea) after the first and the second dose of ChAdOx1 n-COV-19 vaccine. Furthermore, the study conducted by (13) also highlights that the safety of the second dose of the ChAdOx1 n-COV-19 vaccine compared to the first dose varies only by age. Specifically, according to the study by (13), 88% of the 18-55 years’ group reported higher systemic side-effects (headache, fever, myalgia, chills and nausea) after receiving the second dose of the ChAdOx1 n-COV-19 vaccine.

The findings based on the inferential Chi-square test in this study also reveals that there is a substantial variation in the prevalence of the local and systemic side-effects among the younger and the older participants. Specifically, the Chi-square test results show that there is a significant difference in the reported incidences of fatigue, discomfort at site and no symptoms, 48 hours after administration of ChAdOx1 n-COV-19 second dose vaccine among the younger (≤ 40 years) and the older (>40 years) participants. In addition, the multiple logistic regression results also presented significant evidence that showed older vaccine recipients have lower odds of reporting fever, body pain, chills, nausea, myalgia, headache, fatigue, and other symptoms 48 hours after administration of the first dose.

The multiple logistic regression findings indicated that 48 hours after receiving the second dose, younger vaccine recipients were more likely to experience fever, body pain, fatigue and discomfort at site compared to older vaccine recipients. Similarly, using multivariate logistics regression analysis to determine the side-effects associated with ChAdOx1 n-COV-19 vaccine side-effects in Saudi Arabia, the study by Alhazmi (15) found that only fatigue and fever were significant side-effects. Specifically, given that the majority of the participants in the study by (15) were female, the implication is that gender had a significant effect in determining the prevalence of fever and fatigue side-effects after administration of the ChAdOx1 n-COV-19 vaccine. The multiple logistics regression results also depicted that in 48 hours after administration of the first and second dose, the older vaccine (>40) recipients had higher odds of reporting no symptoms compared to younger vaccine recipients. The implication is that younger individuals reported higher frequency of the systemic and local side-effects compared to the older participants. The stated insight is also consistent with the previous findings by Abu-Hammad (16) who noted that among the Jordanian healthcare workers that had received ChAdOx1 n-COV-19 vaccine, only age group but not gender was significantly associated with the severity of side effects after the first dose. However, according to (15), neither age nor gender was significantly associated with the severity of the local side-effects after administration of the second dose. The insight based on the manufacturer’s trial study also indicated the claim that the ChAdOx1 n-COV-19 vaccine is better tolerated in older recipients compared to the younger vaccine recipients with the same immune system (17). This means that older recipients tend to have less local and systemic side-effects compared to the younger vaccine recipients.

The stated outcome of the study on variation in vaccine side-effects by age based on Chi-square and multiple logistic regression analysis is also consistent with the study by (13). According to the stated study, the ChAdOx1 n-COV-19 vaccine seems to be better tolerated in older recipients compared to the younger individuals. Specifically, (13) reports that while individuals, 18-55 years reported 88% incidences of local side-effects, only 61% of those above 70 years complained of experiencing local side-effects associated with the ChAdOx1 n-COV-19 vaccine. Madhi et al. (12) also report that younger recipients of the ChAdOx1 n-COV-19 vaccine are more likely to experience systemic and local side-effects immediately after vaccination. However, these findings contrast the insight based on the study by Hatmal et al. (11) who did not find any notable differences between the rate of systemic and local side-effects among the younger and the older ChAdOx1 n-COV-19 vaccine recipients in Jordan.

The authors of this study found that female respondents are more susceptible to the side-effects compared to the male participants, although the sample composition might have influenced the results. The multiple logistic regression analysis outcome indicated that 48 hours after receiving the first dose, female vaccine recipients had a greater likelihood of complaining of body pain, chills, nausea, headache, fatigue, and discomfort at site compared to male vaccine recipients. The findings also indicated that 48 hours after receiving the second dose, female vaccine recipients also had greater odds of reporting headache, fatigue, and discomfort at site.

### 4.2 Implication of Findings

The findings have important implication on the public health efforts to raise awareness among the public with respect to vaccine hesitancy. The fact that the reported rate of occurrence of the side-effects was mostly less than 50% is consistent with the manufacturer’s claim that the ChAdOx1 n-COV-19 vaccine is safe (17, 18). Furthermore, the insight that the severity and incidences of the reported side-effects decrease 48 hours after the second dose compared to 48 hours after the first dose suggest that the adverse side-effects are mild (19).

## 5.0 Conclusion

The study concludes and acknowledges that the ChAdOx1 n-COV-19 vaccine is safe in consistent with the manufacturer’s claim. The reported prevalence of the systemic side-effects decreases significantly 48 hours after administration of the second dose compared to 48 hours after administration of the first dose. However, older individuals have lower reported incidences of the systemic and local side-effects compared to the younger individuals. The implication is that there is a significant difference in the reported systemic and local side-effects by age of the vaccine recipients. Specifically, the study found that younger vaccine recipients (≤ 40 years) reported higher cases of fatigue and discomfort at site compared to older vaccine recipients (>40 years). The study also found that the reported prevalence of systemic and local side-effects varied significantly by gender (sex) of the vaccine recipients. On average, female vaccine recipients had higher reported incidences of systemic and local side-effects 48 hours after administration of the first and the second dose compared to the male vaccine recipients. Generally, there were more reported systemic side-effects compared to the local side-effects 48 hours after administration of the first and the second dose of the ChAdOx1 n-COV-19 vaccine. The findings have important implication for public health policy to lower vaccine hesitancy. Ethical approval was granted by the Ministry of Health, Trinidad and Tobago (3/13/441 Vol. II) and the North Central Regional Health Authority, Trinidad and Tobago.

## Data Availability

Data is available upon request

## 6.0 Declarations

### 6.2 Authors’ Contributions

CDG and DV were responsible for data analysis, with intellectual contributions from DT. CDG and DV drafted the article. All authors contributed to the conception and design of the paper, interpretation of data, and critical revisions contributing to the intellectual content and approval of the final version of the manuscript.

## References

1. Majeed A, Papaluca M, Molokhia M. Assessing the long-term safety and efficacy of Covid-19 vaccines. Journal of the Royal Society of Medicine. 2021; 1(1):1–10.

2. Majeed A, Molokhia M. Vaccinating the UK against Covid-19. BMJ Journal. 2020; 371(1):4654.

3. WHO. Side-effects of Covid-19 vaccines [internet]. 2021. Available from: https://www.who.int/news-room/feature-stories/detail/side-effects-of-covid-19-vaccines/

4. Razai MS, Osama T, McKechnie DGJ, Majeed A. Covid-19 hesitancy among ethnic minority groups. BMJ Journal. 2021; 372(1):513.

5. Torjesen I. Covid 19: Norway investigates 23 deaths in frail elderly patients after vaccination. BMJ Journal. 2021; 371(1):4780.

6. Riad A, Pokorna A, Attia S, Klugarova J, Koscik M, Klugar M. Prevalence of Covid-19 vaccine side effects among healthcare workers in Czech Republic. Journal of Clinical Medicine. 2021; 10(1428):1–18.

7. Gopaul C, Ventour D, Trotman M, Thomas D. The Epidemiology Characteristics of Positive COVID-19 patients in Trinidad and Tobago. MedRxiv, 2020; [Preprint] Available from: https://doi.org/10.1101/2020.08.06.20148288

8. Gopaul C, Ventour D, Thomas D. Laboratory Predictors for COVID-19 ICU Admissions in Trinidad and Tobago. Research Square, 2020; [Preprint] Available from: https://doi.org/10.21203/rs.3.rs-103394/v1

9. Menni C, Klaser K, May A, Polidori L, Capdevila J, Louca P. Vaccine side-effects and SARS-CoV-2 infection after vaccination in users of the COVID Symptom Study app in the UK: a prospective observational study. The Lancet Infectious Diseases Journal, 2021; 21(7):939–949.

10. Bernal JL, Andrews N, Gower C, Robertson C, Stowe J, Tessier E. Effectiveness of the Pfizer-BioNTech and Oxford-AstraZeneca vaccines on covid-19 related symptoms, hospital admissions, and mortality in older adults in England: test negative case-control study. The BMJ Journal, 2021; 373(1):1–5.

11. Hatmal MM, Al-Hatamleh AI, Olaimat AN, Hatmal M, Alhaj-Qasem DM, Olaimat TM, et al. Side effects and perceptions following COVID-19 vaccination in Jordan: A randomized, cross-sectional study implementing machine learning for predicting severity of side effects. Vaccines, 2021; 9(6):556.

12. Madhi SA, Baillie V, Cutland CL, Voysey M, Koen AL, Fairlie L, et al. Safety and efficacy of the ChAdOxI n-Cov-19 (AZDI 222) Covid-19 vaccine against the B.I..35I variant in South Africa. medRxiv, 2021; 1–26.

13. Ramasamy MN, Minassian AM, Ewer KJ, Flaxman AL, Folegatti PM, Owens DR, et al. Safety and immunogenicity of ChAdOx1 nCoV-19 vaccine administered in a prime-boost regimen in young and old adults (COV002): a single-blind, randomised controlled trial. Lancet Journal, 2021; 396 (1):1979–193.

14. Voysey M, Clemens SAC, Madhi SA, Weckx LY, Folegatti PM, Aley PK, et al. Single-dose administration and the influence of the timing of the booster dose on immunogenicity and efficacy of ChAdOx1 nCoV-19 (AZD1222) vaccine: a pooled analysis of four randomised trials. Lancet Journal, 2021; 397(1):881–891.

15. Alhazmi A, Alamer E, Daws D, Hakami M, Darraj M, Abdelwahab S, et al. Evaluation of Side Effects Associated with COVID-19 Vaccines in Saudi Arabia. Vaccines. 2021; 9(6):674.

16. Abu-Hammad O, Alduraidi H, Abu-Hammad S, Alnazzawi A, Babkair H, Abu-Hammad A, et al. Side Effects Reported by Jordanian Healthcare Workers Who Received COVID-19 Vaccines. Vaccines. 2021; 9(6):577.

17. AstraZeneca. AZD1222 US Phase III trial met primary efficacy endpoint in preventing COVID-19 at interim analysis[online]. 2021. Available from: https://www.astrazeneca.com/

18. Walsh EE, Frenck RW, Falsey AR, Kitchin N, Absalon J, Gurtman A, et al. Safety and immunogenicity of two RNA-based COVID-19 vaccine candidates. The New England Journal of Medicine. 2020; 383(1):2439–2450.

19. Hazell L, Shakri SAW. Under-reporting of adverse drug reactions. A systematic review. Drug Safety, 2006; 29 (1):385–396.

